# A prospective cohort study of the clinical profile of *Acinetobacter baumannii* infections in Thailand

**DOI:** 10.64898/2026.03.26.26349299

**Authors:** Saranta Freeouf, Samantha Palethorpe, Cassandra Fairhead, Chidchamai Kewcharoenwong, Supphachoke Khemla, Nopparat Wiboonsunti, Sangravee Juhong, Brendan Wren, Tansy Edwards, Ganjana Lertmemongkolchai, Jeremy Brown

## Abstract

**Objectives:** To better define the clinical features of *Acinetobacter* spp. infection in Northern Thailand, including a comparison of hospital- and community-acquired infections (HAIs and CAIs).

**Methods:** A prospective clinical study of *Acinetobacter* spp. infections at two Northern Thailand hospitals from 2019 to 2022, collecting data on sample sources, patient demographics, comorbidities, antimicrobial resistance profiles, and outcomes.

**Results:** Of 129 enrolled patients, 81.4% had *Acinetobacter* spp. isolated from a respiratory sample. A significant minority (25.6%) of infections were CAIs, 33.3% of which were admitted to ITU within 24 hours of admission. Compared to HAIs, CAIs were significantly more likely to be caused by blood (15.2%, *p*=0.0258), wound (21.2%, *p*=0.0120), or urine infections (12.1%, *p*=0.0370). *Acinetobacter* spp. HAIs mainly occurred after admission to ITU (87.7%, *p*<0.0001) and were more likely to be multidrug-resistant than CAIs (76.3% vs. 34.4%, *p*<0.0001). Overall, the median length of hospital stay was 27 days and there was a 27.1% in-hospital mortality, which was increased in patients with CVA/brain (*p*=0.005), and multidrug-resistant (*p=*0.010) or carbapenem-resistant infections (*p=*0.003).

**Conclusions:** These data define the clinical profile of *Acinetobacter* spp. infections in Northern Thailand, confirming their high mortality and demonstrating CAIs are a significant proportion of all cases.

## Introduction

Antimicrobial (AMR) resistant bacterial infections are described by the World Health Organisation (WHO) as a major global health challenge, estimated to be responsible for nearly 5 million deaths in 2019 (1) and predicted to rise to 10 million deaths yearly by 2050 (2). The Gram-negative bacterial pathogen *Acinetobacter baumannii* has particularly high levels of antimicrobial resistance and is the fifth commonest AMR bacterial cause of death globally (3). The majority of global deaths associated with *A. baumannii* infection are due to pneumonia or septicaemia, although it also causes a wide range of other infections including wound, urinary tract, and indwelling device infections and meningitis (3). AMR *A. baumannii* infections are particularly prevalent in Southeast Asian countries, including Thailand where they cause over an estimated 15,000 deaths per year (4)*. A. baumannii* is a major cause of hospital-acquired infection (HAI); for example, Thai data shows *Acinetobacter* spp. caused 16% of HAI septicaemias (5) and carbapenem-resistant *A. baumannii* (CRAB) isolates were responsible for 4.5% of all HAIs (6). More recent data show CRAB caused 51% of all AMR bacterial hospital-acquired septicaemias across Thailand (7). Similarly, a multi-national study from 10 Asian countries showed *Acinetobacter* spp. caused 14% of hospital-acquired pneumonia (HAP) cases and 37% of ventilator-acquired pneumonia (VAP) cases, and these were associated with high antibiotic resistance rates (8). Although isolation of *A. baumannii* from respiratory samples from ventilated patients could reflect colonisation rather than a true VAP, the attributable mortality of *A. baumannii* has been calculated to be 10% to 43% for intensive therapy unit (ITU) patients and 7.8% to 23.0% for general ward patients (9), indicating that *A. baumannii* was likely to be causing active infection in a high proportion of cases.

As well as causing HAIs, *A. baumannii* has been reported to cause community-acquired infections (CAIs), mainly pneumonia that is often associated with comorbidities, heavy smoking, and excess alcohol consumption (10,11). Although *A. baumannii* was previously considered a relatively rare cause of CAIs (10,12,13), recent publications show it is a common cause of community-acquired pneumonia (CAP) in Nepal, Indonesia, Sri Lanka and China (14–19) and a major cause of community-acquired septicaemia in Taiwan, China, and Thailand (6,20–22). Overall, the existing data suggest that globally *A. baumannii* mainly causes HAIs, but in certain geographic locations it is also an important cause of CAIs. *A. baumannii* is the main pathogenic *Acinetobacter* species, but other *Acinetobacter* spp. can also cause serious infections and can be incorrectly identified as *A. baumannii* by standard microbiological techniques. For example, recent data from Thailand showed that 29% of reported *A. baumannii* infections were actually caused by *A. nosocomialis* (23), and these infections also had high rates of antibiotic resistance and mortality (33% and 36%, respectively).

Despite its clinical importance, there are a paucity of detailed prospectively collected clinical data on *Acinetobacter* spp. infections. Detailed data on whether *Acinetobacter* spp. infections are hospital- or community-acquired, patient demographics, the clinical syndromes commonly caused by *Acinetobacter* spp. infections, and subsequent mortality are essential for the design of clinical trials of new antibiotic regimens (24) and therapeutic approaches such as monoclonal antibodies (25,26). To help fill this gap, we have conducted a prospective cohort study of patients presenting with microbiologically-proven *A. baumannii* and *A. nosocomialis* infections at two Thai provincial hospitals. We aimed to define the overall clinical spectrum and outcomes of *A. baumannii* and *A. nosocomialis* infections, quantify the relative proportions of CAIs versus HAIs, and assess any differences in the clinical presentation between CAI and HAI cases.

## Methods

### Study design, participants, and setting

A prospective cohort study designed to evaluate the clinical profiles and outcomes of HAI and CAI infections caused by *A. baumannii* and *A. nosocomialis* in two secondary care hospitals in Northern Thailand between 2019 and 2022. Patients were prospectively recruited from Lamphun Hospital, a secondary care hospital in Lamphun province, Northern Thailand from 22nd July 2021 to 11^th^ August 2022, and Nakhon Phanom Hospital, a secondary care hospital in Nakhon Phanom province, Northeastern Thailand from 8^th^ April 2019 to 30^th^ June 2021 (with pauses during COVID lockdowns). All consenting inpatient adults with a positive culture for *Acinetobacter* spp. initially identified by the local microbiology services as *A. baumannii* from any sample were eligible for inclusion. Written informed consent was obtained prior to enrolment (study ethical approval: NP-EC11-No.1/2562 and LPN52/2563). Cases were defined as CAI if they were culture positive within 72 hours of their admission date, unless transferred from another health centre with a different admission diagnosis, and HAI if culture positive >72 hours after admission or transferred from other health centres where they had a different admission diagnosis.

### Clinical data and microbiology

The local research team obtained data from hospital records on age, sex, comorbidities, smoking history, season of admission (Summer: 16th February to 15th May, Rainy: 16th May to 15th October, Winter: 16th October to 15th February), admission diagnosis for HAI cases, isolate antibiotic sensitivities, other positive microbiology isolated on the same day as isolation of *Acinetobacter* spp., admission to ITU at time of isolation, changes in antibiotic use after isolation of *Acinetobacter* spp., duration of hospital stay, and inpatient mortality. Mortality within 90 days of hospital discharge was collected where available; however, many patients were lost to follow up after discharge and hence 90 day mortality rates represent a minimum mortality. Sample sources were divided into the following categories: respiratory (sputum, bronchoscopy samples, or for intubated patients endotracheal aspirates), blood, urine, or pus from any wound sites. For patients with a positive respiratory sample, available chest X-rays were reviewed by an experienced respiratory physician (JSB) and categorised into either showing consolidation or other active infective changes, no radiological evidence of active infection, or indeterminate changes not clearly suggestive of new infection or showing limited change compared to X-rays prior to *Acinetobacter* spp. isolation. *Acinetobacter* spp. were isolated via culture, with identities initially confirmed using the BD Phoenix M50 system at Nakhon Phanom Hospital and the Microscan DxM1040 at Lamphun Hospital. Local reporting policies dictate that all isolates within the *A. baumannii* complex are reported as *A. baumannii*. Antibiotic resistance rates were established at both centres using their available antibiotic panels. To confirm the *Acinetobacter* species, isolates were further characterised at the Department of Medical Technology, Faculty of Associated Medical Sciences, Chiang Mai University, using an established internal PCR method to differentiate between *A. baumannii*, *A. nosocomialis*, and other *Acinetobacter* spp. (23).

### Statistical analysis

Where applicable, data were analysed using GraphPad Prism version 10.1.0 (316) to calculate *p* values using Fisher’s exact test, or two-way ANOVAs, as indicated in legends.

## Results

### Patient recruitment, demographics, and admission characteristics

Within the recruitment periods for both hospital sites *Acinetobacter* spp. were isolated from 612 admissions, of which 141 individuals were initially enrolled to this study (**Fig. 1a**). On further review, twelve cases were excluded because they were infected with an *Acinetobacter* spp. other than *A. baumannii* or *A. nosocomialis*. Of the remaining 129 cases, 97 (75%) were recruited from Nakhon Phanom Hospital and 32 (25%) from Lamphun Hospital (**Table 1**). Patients were recruited at Nakhon Phanom between April 2019 and June 2021, representing 20.0% of all cultures reported by the local microbiology services as positive for *Acinetobacter* spp. at that site for that period (**Supplementary Table 1**). The majority (26/32, 81.3%) of patients at Lamphun Hospital were recruited between July 2021 and October 2021, representing 25.4% of patients with a positive culture for *Acinetobacter* spp. over that period (**Supplementary Table 1**); the remaining patients were recruited intermittently up to August 2022. For Nakhon Phanom, the highest rate of cases was during the rainy season (0.77 per day) with similar rates for winter and summer seasons (0.52 and 0.50 per day respectively), and this was reflected in the seasonal distribution of recruited cases (0.09, 0.11, and 0.15 recruits/day for summer, winter, and rainy seasons respectively) (**Fig. 1b**). The short recruitment period at the Lamphun site prevented assessing for any seasonal effect. Patient characteristics, microbiological data, and outcomes for recruited cases are shown in **Table 1**. All individuals were Thai nationals, 59.7% were male and the median age was 66 years. The majority of *Acinetobacter* spp. isolates (81.4%) were obtained from respiratory cultures (*p* = 0.0004), with smaller numbers of isolates from blood culture, urine, and, for the Nakhon Phanom site only, wound pus (mainly from lower limb lesions, data not shown) (**Table 1**). This infection source distribution was broadly similar for all cases of *Acinetobacter* spp. identifications over the recruitment period at both hospital sites, although these also included additional rarer sites of infection (e.g. peritoneal dialysis or pleural fluid) (**Fig. 1c & d, Supplementary Table 1**).

**Figure 1.**
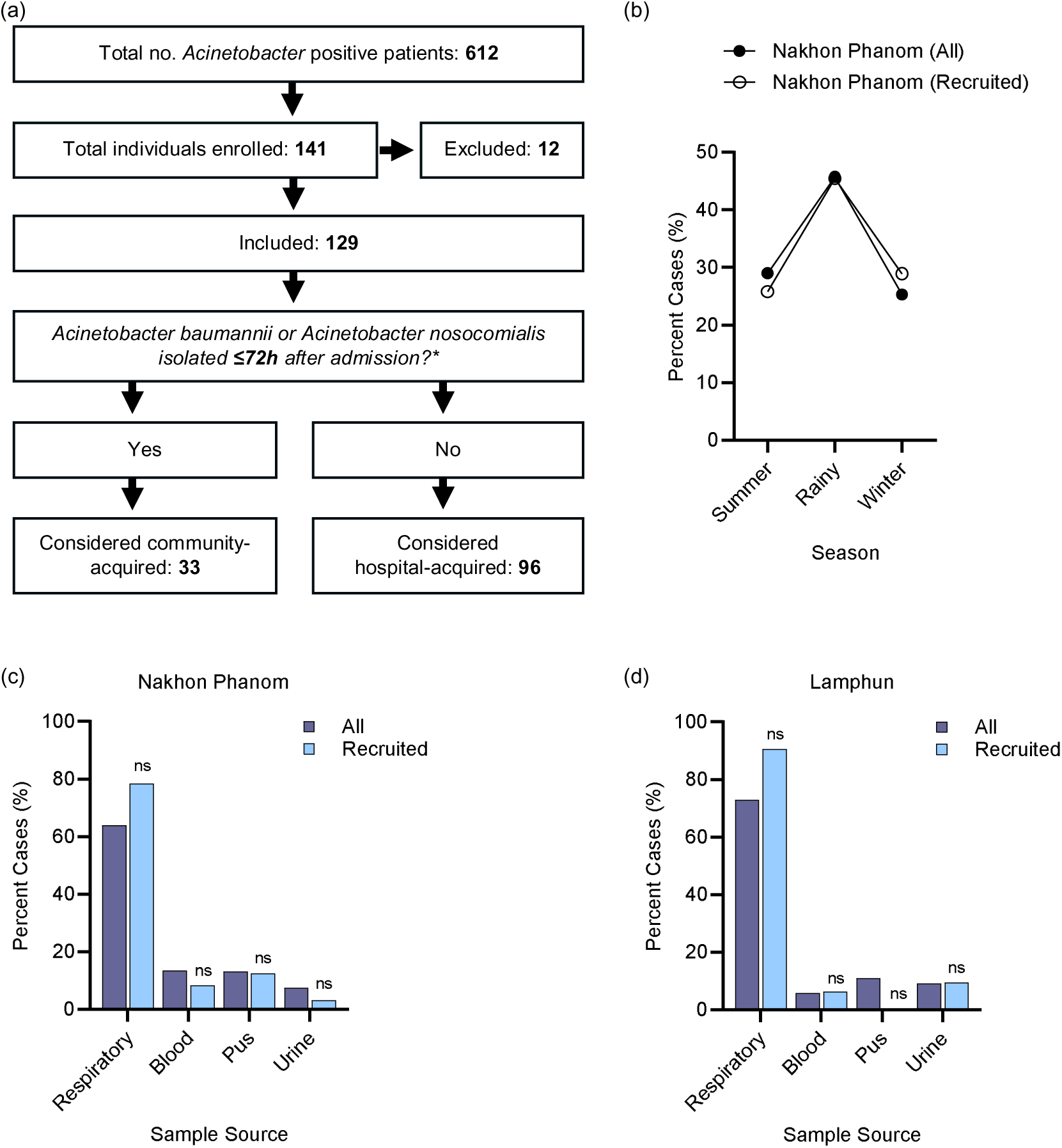
Admission characteristics of patients infected with *Acinetobacter* spp. **(a)** Enrolment flowchart; reasons for exclusion included isolation prior to admission, no sample source data, or *Acinetobacter* spp. unknown or confirmed *Acinetobacter variabilis.* *Considered HAI if culture positive >72h after admission OR transferred to hospital from another health centre with a different admission diagnosis to the *Acinetobacter* spp. infection. **(b)** Percent of all admissions versus recruited admissions diagnosed with *Acinetobacter* spp. infections at Nakhon Phanom during the Summer (16^th^ February – 15^th^ May), Rainy (16^th^ May – 15^th^ October), and Winter seasons (16^th^ October – 15^th^ December) for the Nakhon Phanom site. **(c & d)** *Acinetobacter spp.* sample collection sites from all patients versus recruited patients admitted to Nakhon Phanom **(c)** and Lamphun **(d)**. Respiratory samples included sputum cultures, bronchoalveolar lavage and endotracheal tube aspirate samples. Data were analysed using two-way ANOVA to compare all vs recruited patient samples **(c & d)** (\**p* < 0.05, \*\**p* < 0.01, \*\*\**p* < 0.001, \*\*\*\**p* < 0.0001, ns; not significant).

**Table 1:** Demographics and admission characteristics of patients infected with *Acinetobacter* spp. Figures in brackets are the % of the total for that column except where stated otherwise.

| Parameter |  | All<br>N = 129 (%) | Hospital site |  |
| --- | --- | --- | --- | --- |
|  |  |  | Nakhon Phanom<br>N = 97 (75%) | Lamphun<br>N = 32 (25%) |
| Demographic factors |  |  |  |  |
| Median age (range) |  | 66 (18 to 90) | 63 (18 to 90) | 69 (47 to 90) |
| Sex | Female | 52 (40.3) | 37 (38.1) | 15 (46.9) |
|  | Male | 77 (59.7) | 60 (61.9) | 17 (53.1) |
| Smoking status | Never | 111 (86.0) | 87 (89.7) | 24 (75.0) |
|  | Ex- or active smoker | 17 (14.0) | 10 (10.3) | 7 (21.9) |
|  | Unknown | 1 (0.01) | 0 (0.0) | 1 (3.1) |
| Microbiology |  |  |  |  |
| CAI<sup>1</sup> or HAI<sup>2</sup> | CAI | 33 (25.6) | 25 (25.8) | 8 (25.0) |
|  | HAI | 96 (74.4) | 72 (74.2) | 24 (75.0) |
| Sample source | Respiratory<sup>3</sup> | 105 (81.4) | 76 (78.3) | 29 (90.6) |
|  | Blood<sup>3</sup> | 8 (6.2) | 6 (8.2) | 2 (6.3) |
|  | Urine<sup>3</sup> | 6 (4.7) | 3 (3.1) | 3 (9.4) |
|  | Wound site / pus | 12 (9.3) | 12 (12.4) | 0 (0.0) |
| *Acinetobacter* spp. | *A. baumannii* | 81 (62.8) | 51 (52.6) | 30 (93.8) |
|  | *A. nosocomialis* | 17 (13.2) | 17 (17.5) | 0 (0.0) |
|  | Unclear | 31 (24.0) | 29 (29.9) | 2 (6.3) |
| Co-isolated bacteria | Yes | 33 (25.6) | 31 (32.0) | 2 (6.3) |
|  | No | 96 (74.4) | 66 (68.0) | 30 (93.7) |
| Outcomes |  |  |  |  |
| Median LOS in days (range) |  | 27 (1 to 577) | 30 (5-577) | 24 (1-139) |
| ITU admission | Overall | 78 (60.5) | 56 (57.7) | 22 (68.8) |
|  | Pre-*Acinetobacter* spp.<sup>4</sup> | 50 (39.4) | 38 (40.9) | 12 (37.5) |
|  | Post-*Acinetobacter* spp.<sup>4</sup> | 28 (22.0) | 18 (18.9) | 10 (31.3) |
| Mortality | In-hospital mortality | 35 (27.1) | 23 (23.7) | 12 (37.5) |
|  | 90 day mortality<sup>5</sup> | 51 (39.5) | 38 (39.2) | 13 (40.6) |
| Median days since culture positive to death (range) |  | 16.5 (1 to 94) | 16 (1 to 84) | 17 (3 to 94) |

<sup>1</sup>CAI: culture positive < 72h after admission
<sup>2</sup>HAI: culture positive >72 h after admission
<sup>3</sup>Includes two cases of dual isolation (one blood and sputum, one urine and sputum) at Lamphun Hospital
<sup>4</sup>Divided into whether the *Acinetobacter* spp. was isolated before or after admission to ITU (excludes two cases when date of ITU admission was unknown)
<sup>5</sup>Minimum value due to patients lost to follow-up

### Acinetobacter spp. sample sources and comparison of A. baumannii and A. nosocomialis

PCR analysis (23) of 30 of the 32 Lamphun isolates confirmed all were *A. baumannii*, whereas 17 of the 68 *Acinetobacter* spp. tested from Nakhon Phanom were identified as *A. nosocomialis* strains with the remaining strains confirmed to be *A. baumannii* (**Table 1**). **Table 2** shows the comparison of the demographic and clinical data for Nakhon Phanom patients infected with *A. nosocomialis* versus those infected with *A. baumannii*. Power was low to detect evidence of differences, although compared to patients with *A. baumannii*, patients with positive cultures for *A. nosocomialis* were more likely to be female (33.3% versus 52.9%), have a respiratory source of infection (78.4% versus 94.1%), and require admission to ITU after isolation of *Acinetobacter* spp. (13.7% versus 29.4%) (**Fig. 2a**, **Table 2**). *A. nosocomialis* infections were also associated with a lower in hospital mortality (23.5% versus 17.6%), but a similar 90 day mortality (39.2% versus 35.3%) (**Fig. 2a**, **Table 2**).

**Table 2:**
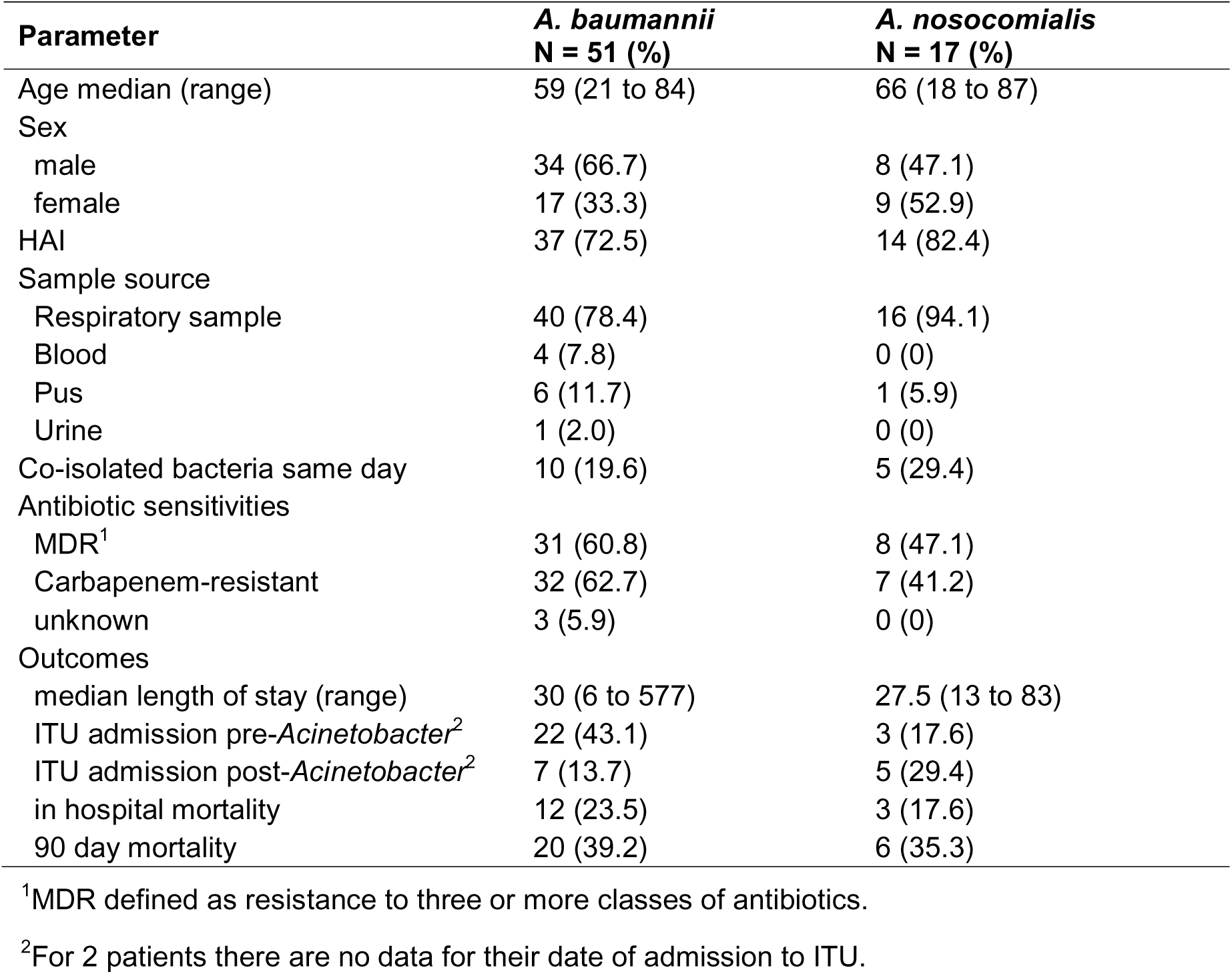
Comparison of data for patients infected with *A. baumannii* to those with *A. nosocomialis*. Data restricted to patients from Nakhon Phanom site with confirmed specification using multiplex PCR (**23**).

**Figure 2.**
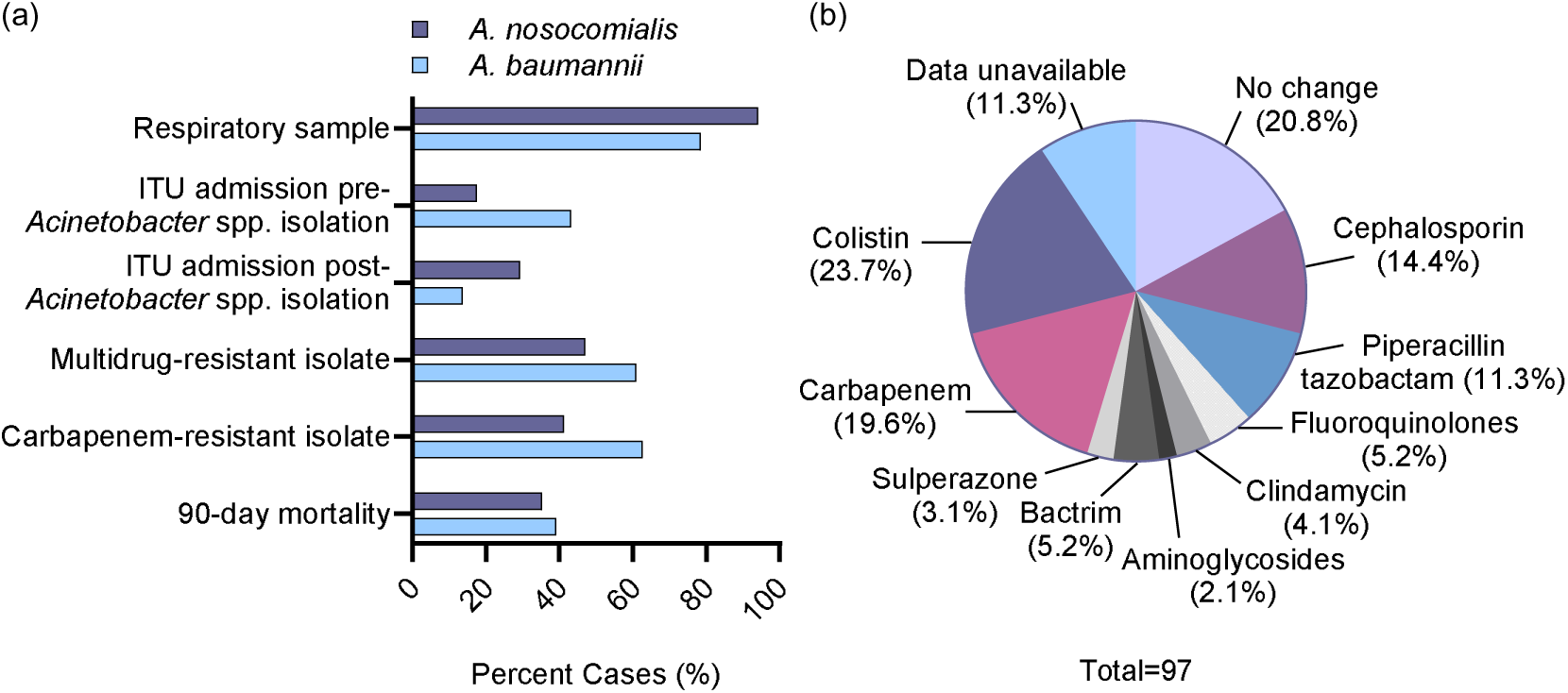
Clinical characteristics of *Acinetobacter* spp. infections. **(a)** Comparative selected clinical data for patients from Nakhon Phanom infected with *A. nosocomialis* versus *A. baumannii*. **(b)** Additional new antibiotics prescribed within 144 hours of *Acinetobacter* spp. isolation from patients in Nakhon Phanom hospital.

### Antibiotic resistance data for Acinetobacter spp. isolates

As expected, across both sites the *Acinetobacter* spp. isolates had high levels of antibiotic resistance. MDR and carbapenem resistance were detected for 82 (65.6%, data available for 125 strains) and 78 (62.9%, data available for 124 strains) strains respectively, and there were high levels of resistance to multiple other antibiotics as well (**Table 3**). When defined as MICs of ≥4 μg/ml, resistance to colistin remained relatively rare (3 of 85 isolates tested, 3.5%) (**Table 3**). Rates of MDR and carbapenem resistance for *A. nosocomialis* isolates were lower than for *A. baumannii* but were still high at 47.1% and 41.2% respectively (**Fig. 2a**, **Table 2**). Isolation of *Acinetobacter* spp. was associated with a change in antibiotic regimen within 144 hours in 79.2% of patients (data only available for Nakhon Phanom site); this change included starting treatment with intravenous colistin in 23.7% of cases (**Fig. 2b, Supplementary Table 2**).

**Table 3:**
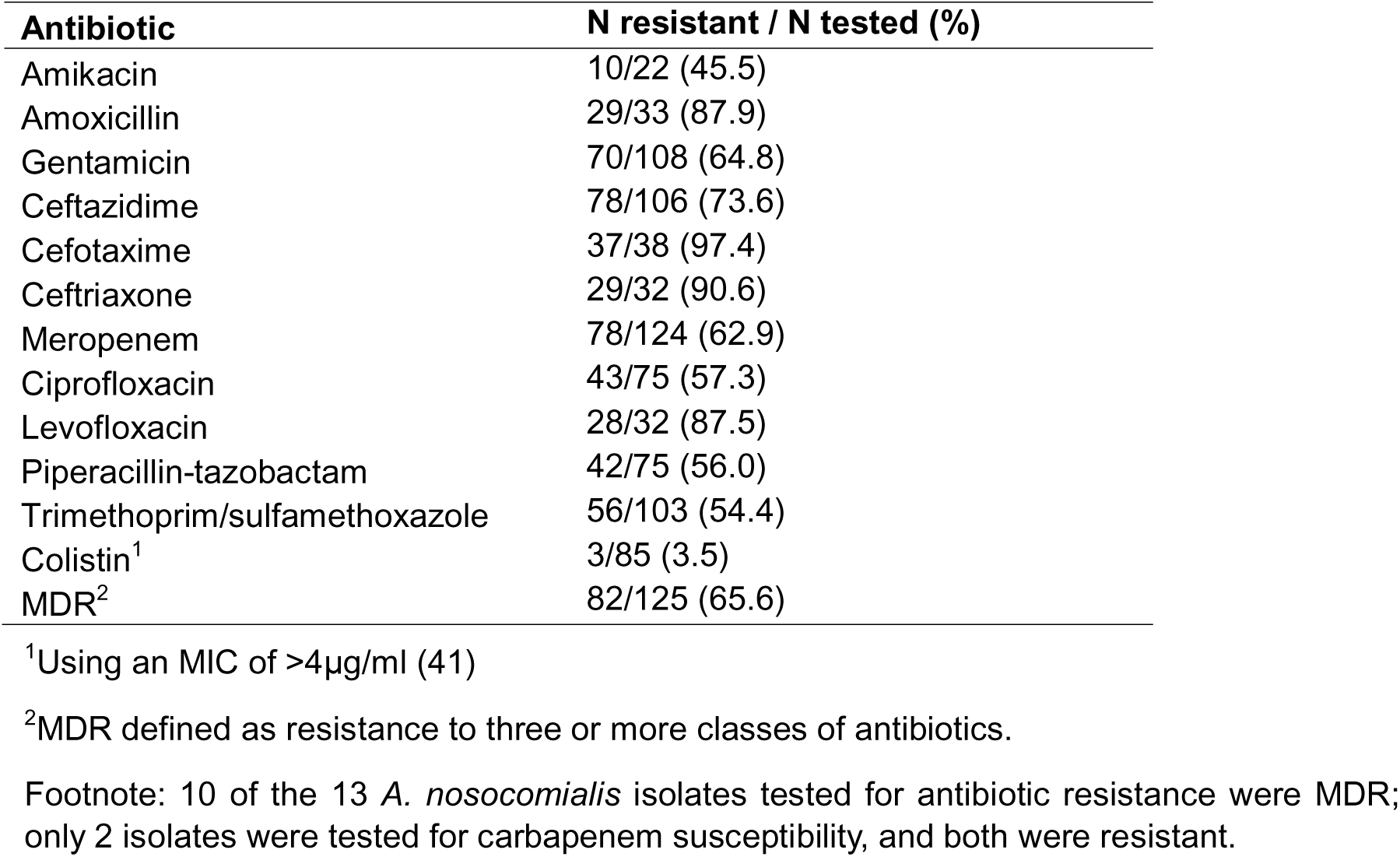
Antibiotic resistance patterns for *Acinetobacter* spp. Isolates.

### Coinfection rates with other bacterial pathogens

Additional potential bacterial pathogens were isolated from the same sample site on the same day as *Acinetobacter* spp. for 33 (25.8%) cases (**Table 4**), the majority of which were recruited from the Nakhon Phanom site (31, 93.9% of cases with coinfections) (**Table 1**). Co-infection rates were similar for blood, pus, and respiratory samples, but no urine samples were positive for additional bacterial pathogens at the time of isolation of *Acinetobacter* spp. (**Table 4**). Co-infections were caused by a range of Gram-positive and Gram-negative pathogens; the commonest were *Escherichia coli, Klebsiella pneumoniae*, *Enterobacter* spp., and *Pseudomonas aeruginosa*. Isolation of two or more additional pathogens occurred in 8 (6.3%) cases, including 33.3% of patients with a positive culture for *Acinetobacter* spp. from a pus sample.

**Table 4:** Potentially pathogenic bacteria co-isolated with *Acinetobacter* spp. from the same sample source on the same day.

| Co-isolated bacteria | *Acinetobacter* spp. sample source |  |  |
| --- | --- | --- | --- |
|  | Blood<br>N = 8 (%) | Pus<br>N = 12 (%) | Respiratory<br>N = 105 (%) |
| None | 5 (62.5) | 8 (66.7) | 79 (75.2) |
| Single extra species | 2 (25.0) | 0 | 23 (21.9) |
| Multiple extra species | 1 (12.5) | 4 (33.3) | 3 (2.9) |
| Species isolated: |  |  |  |
| *Escherichia coli* | 2 (25.0) | 3 (25.0) | 1 (1.0) |
| *Klebsiella pneumoniae* | 0 | 1 (8.3) | 4 (3.8) |
| *Stenotrophomonas maltophilia* | 0 | 2 (16.7) | 2 (1.9) |
| *Enterobacter* spp. | 0 | 2 (16.7) | 3 (2.9) |
| *Enterococcus faecalis* | 0 | 1 (8.3) | 1 (1.0) |
| *Pseudomonas aeruginosa* | 0 | 1 (8.3) | 4 (3.8) |
| *Staphylococcus aureus* | 0 | 0 | 4 (3.8) |
| Gram positive cocci | 1 (12.5) | 0 | 1 (1.0) |
| *Proteus mirabilis* | 0 | 0 | 1 (1.0) |
| *Haemophilus influenzae* | 0 | 0 | 1 (1.0) |

In addition, 4 subjects with *Acinetobacter* spp. in their respiratory sample had positive blood cultures on the same day for an additional pathogen (2 *S. aureus*, 2 *K. pneumoniae*)

### Comparison of CAI and HAI Acinetobacter spp. cases

CAI accounted for 33 (25.6%) and HAI for 96 (74.4%) of *Acinetobacter* spp. cases (**Table 1**). For HAI cases, the original admission to hospital was for a wide range of clinical conditions, with other infections and CNS disease the commonest reasons (**Fig. 3a, Supplementary Table 3**). **Table 5** compares the clinical parameters for CAI versus HAI cases. There were no apparent differences in median age, sex ratio, hospital site, or co-morbidities between HAI and CAI cases. More CAI patients were admitted during the summer season compared to HAI cases, but for both categories the highest proportion of cases were admitted during the rainy season (**Fig. 3b**). *A. nosocomialis* caused both CAI and HAI cases (**Table 5**). Sample sources did differ between CAI and HAI cases, with HAI isolates predominantly coming from respiratory samples (89.6% vs 57.6% for CAI, *p* < 0.001), whereas a higher proportion of CAI cases had a positive culture from blood (*p* = 0.026), pus (*p* = 0.012), or urine (*p* = 0.037) compared to HAIs (**Fig. 3c, Table 5**). There was evidence of lower rates of MDR or carbapenem resistance in CAI *Acinetobacter* spp. isolates compared to HAI cases (*p* < 0.001 for both), although these were still high at 34.4% and 32.2% respectively (**Figure 3d, Table 5**). Chest X-rays taken within 48 hours of isolation of *Acinetobacter* spp. from a respiratory sample showed radiological evidence of active infection for 14 of 19 (73.7%) CAI cases with an available chest X-ray, suggesting community acquired pneumonia was the diagnosis (**Fig. 3e, Supplementary Table 4**). For HAI cases with *Acinetobacter* spp. isolated from respiratory samples, a substantially lower proportion of cases was associated with clear infective changes on the chest X ray (31.4%, *p* = 0.001) (**Fig. 3f, Supplementary Table 4**). Instead, a higher proportion had radiological abnormalities which did not clearly represent new infective infiltrates (33.7%, *p* = 0.0117) and a 22.1% of cases had no radiological evidence of active lung infection; a proportion of these patients may represent colonisation of the respiratory tract with *Acinetobacter* spp. rather than active infection. Of HAI cases 50 (52.8%) were already admitted to ITU for a median duration of 14.5 days prior to isolation of *Acinetobacter* spp.; of these, *Acinetobacter* spp. were isolated from a respiratory sample in 90%. Over half of CAI patients were admitted to ITU (**Table 5**), including 11 (33.3%) on the same day of admission; a much higher rate than for non-ITU patients developing HAI with *Acinetobacter* spp. (56.3% versus 10.4%, *p* < 0.001). Despite the high rates of admission to ITU, the 90 day mortality of CAI was lower than for HAI (30.3% versus 49.0%, *p* = 0.071) (**Table 5**).

**Figure 3.**
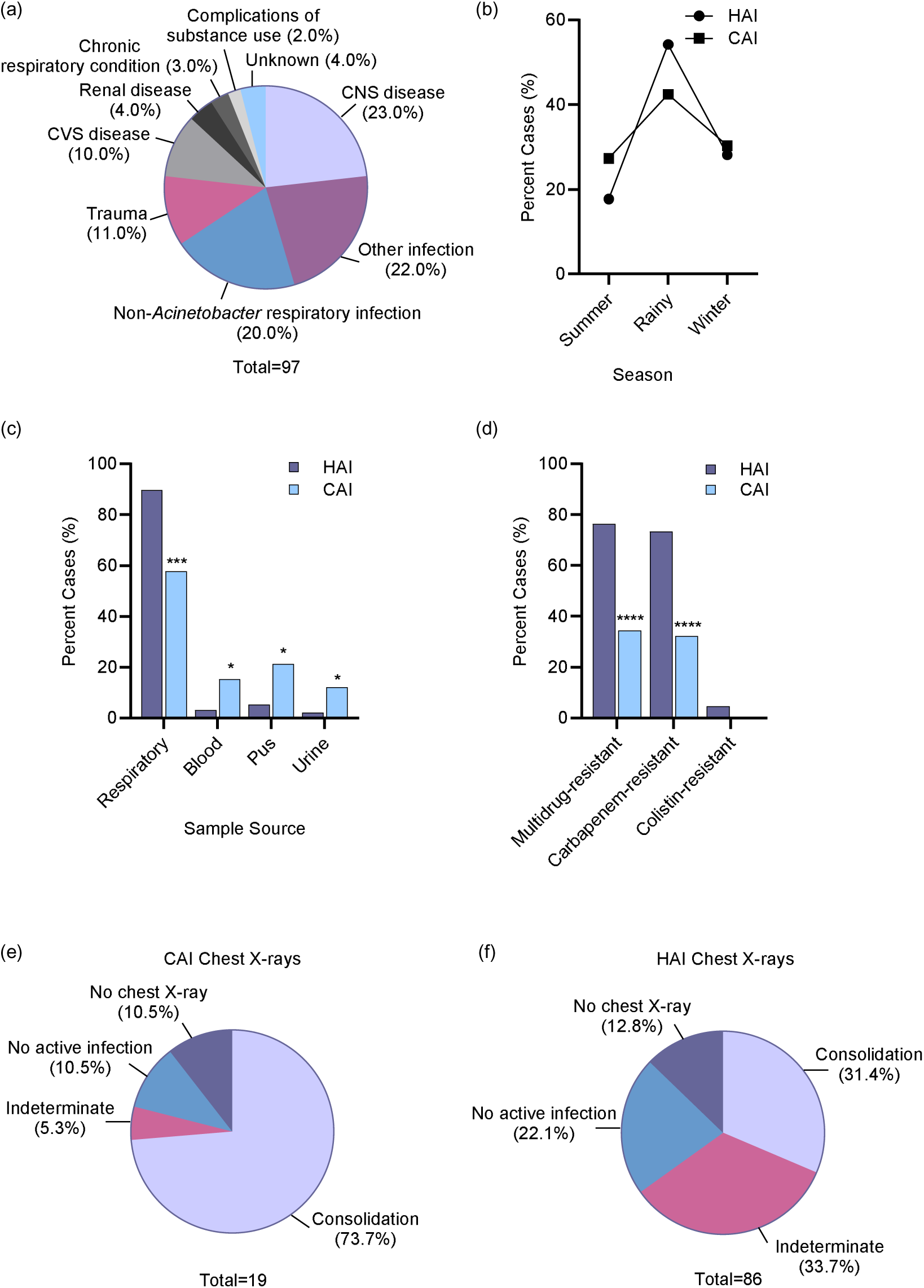
Clinical characteristics of hospital-acquired (HAI) versus community-acquired (CAI) *Acinetobacter* spp. infections. **(a)** Primary reasons for admission for HAI cases. **(b)** Proportion of HAI and CAI cases diagnosed during the Summer (16^th^ February – 15^th^ May), Rainy (16^th^ May – 15^th^ October), and Winter seasons (16^th^ October – 15^th^ December). **(c)** Sites of isolation of *Acinetobacter spp.* from patients with HAI and CAI (respiratory samples included sputum cultures, bronchoalveolar lavage and endotracheal tube aspirate samples). **(d)** Antibiotic resistance rates for HAI and CAI *Acinetobacter* spp. isolates (multidrug-resistant = resistant to three or more classes of antibiotics). **(e & f)** Chest X-ray appearances in CAI **(e)** and HAI **(f)** cases. For **(c)** and **(d)** comparisons between HAI and CAI data were analysed using Fisher’s exact test (\**p* < 0.05, \*\**p* < 0.01, \*\*\**p* < 0.001, \*\*\*\**p* < 0.0001, ns; not significant).

**Table 5:** Comparison of clinical parameters for CAI versus HAI *Acinetobacter* spp. infection.

| Parameter | CAI<br>N = 33 (%) | HAI<sup>5</sup><br>N = 96 (%) |
| --- | --- | --- |
| Hospital site |  |  |
| Lamphun | 8 (24) | 24 (25) |
| Nakhon Phanom | 25 (76) | 72 (75) |
| Age median (IQR) | 66.0 (51.0-71.0) | 65.5 (55.0-74.3) |
| Sex |  |  |
| male | 21 (63.6) | 56 (58.3) |
| female | 12 (36.4) | 40 (41.7) |
| *A. nosocomialis* | 3 (9.1) | 14 (14.5) |
| Smoking status | 5 (15.2) | 12 (12.5) |
| Comorbidities |  |  |
| diabetes | 13 (39.4) | 28 (29.2) |
| renal disease | 8 (24.2) | 17 (17.7) |
| CVA / chronic neurological | 8 (24.2) | 30 (31.4) |
| hypertension | 12 (36.4) | 39 (40.6) |
| chronic lung disease | 3 (9.1) | 12 (12.5) |
| neoplasia | 0 (0) | 2 (2.1) |
| Sample source<sup>1</sup> |  |  |
| Respiratory sample | 19<sup>1</sup> (57.6) | 86 (89.6)*** |
| Blood | 5<sup>1</sup> (15.2) | 3 (3.1)* |
| Pus | 7 (21.2) | 5 (5.2)* |
| Urine | 4<sup>1</sup> (12.1) | 2 (2.1)* |
| Co-isolated bacteria | 11 (33.3) | 22 (22.9) |
| Antibiotic sensitivities |  |  |
| Multidrug-resistant<sup>2</sup> | 11/32 (34.4) | 71/93 (76.3)**** |
| Carbapenem-resistant | 10/31 (32.2) | 68/93 (73.3)**** |
| Colistin- resistant | 0/18 (0) | 3/67 (4.5) |
| Outcomes |  |  |
| median length of stay<sup>3</sup> (range) | 15 (1 to 577) | 32 (7 to 355) |
| post-*Acinetobacter* ITU<sup>4</sup> | 18/32 (56.3) | 10/81 (12.3)**** |
| in hospital mortality | 6 (18.2) | 29 (30.2) |
| 90 day mortality | 10 (30.3) | 47 (49.0) |

<sup>1</sup>Includes two cases of dual isolation (one blood and sputum, one urine and sputum) at Lamphun Hospital
<sup>2</sup>MDR defined as resistance to three or more classes of antibiotics
<sup>3</sup>Data missing for LOS for 1 HAI and 1 CAI patient
<sup>4</sup>ITU timing data missing for 2 HAI cases
<sup>5</sup>Statistical significance determined by Fisher's exact test (\* $p < 0.05$ , \*\* $p < 0.01$ , \*\*\* $p < 0.001$ , \*\*\*\* $p < 0.0001$ )

### Associations with mortality

The median length of hospital stay for patients with *Acinetobacter* spp. was 27 days (range 1-577) and mortality was high (27.1% in-hospital mortality, increasing to at least 39.5% at 90 days). The median time to death from isolation of *Acinetobacter* spp. was 16.5 days (range 1-94), (**Table 1**). There was no evidence that age, sex, co-morbidities of diabetes, renal disease, smoker, chronic lung condition, or hypertension, HAIs, admission to ITU, and co-isolated bacteria were associated with mortality (**Table 6**). However, patients with CVA/brain (*p* = 0.005), infected with an MDR strain (*p* = 0.010) or CRAB strain (*p* = 0.003), and possibly a respiratory source of infection (*p* = 0.081) had higher odds of mortality (**Table 6**).

**Table 6:**
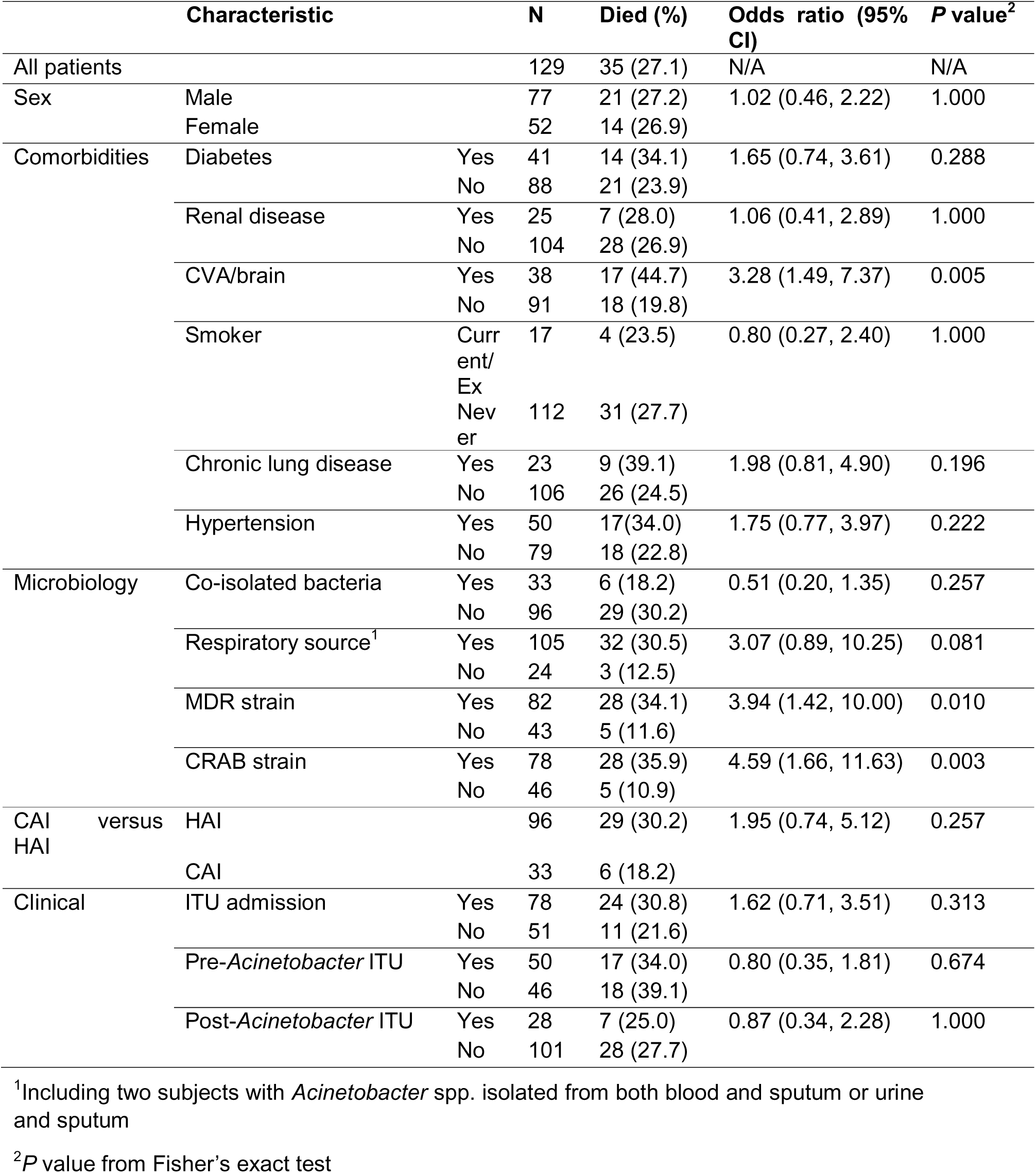
In-patient mortality related to selected clinical and microbiological factors.

## Discussion

The rapid rise in the incidence of infections caused by AMR *Acinetobacter* spp. has emphasised their increasing clinical importance, yet there have only been a limited number of prospective studies assessing the clinical picture of infections associated with *Acinetobacter* spp. Furthermore, there seems to be considerable geographical variation in the nature of *Acinetobacter* spp. infections, which in Western countries remain largely restricted to HAIs mainly affecting ITU patients (10,27–29), whereas in Asia *A. baumannii* (and perhaps other *Acinetobacter* species) are common causes of HAIs both on general wards and in ITU (4–7), and also are a cause of CAIs (6,18,20–22). In this context, we have performed a prospective study at two general hospitals in Thailand to better define the clinical spectrum of *Acinetobacter* spp. infections in a country with a high incidence of infection. Our data has confirmed *Acinetobacter* spp. infections are common, affecting approximately 2 and 4 patients per week at each of our hospital sites. More detailed analysis of 129 cases showed that similar to previous Asian studies our patients with *Acinetobacter* spp. infections had a median age of 66 years with a male predominance, and a substantial level of comorbidities (5,18,30–32). The respiratory tract was by far the commonest site of infection, with wound pus the second commonest source, and as expected AMR rates were very high. Important additional findings were (i) infections were commoner in the rainy season (16th May to 15th October), (ii) there were high rates of co-infection with other potential pathogens, and (iii) CAI infections represented a quarter of cases at both sites which is a similar figure to the results of small retrospective Thai studies (4,20,33) but much higher than larger global studies (30). As previously reported (4,9,23), overall outcomes were poor with a hospital mortality of 27%. However, even with incomplete data our 90 day mortality was significantly higher at 39.5%. Along with CVA/brain injury, in hospital mortality was associated with infection with an MDR or CRAB strain and likely with a respiratory source of infection all of which were commoner in HAI compared to CAI cases. However, although HAI cases had a higher mortality than CAI, we did not have power to detect this in our cohort. Similarly, we did not detect an increase in mortality for patients admitted to ITU during their admission, although this would be a plausible hypothesis in a much larger cohort. Our data has also emphasised the high burden on medical services caused by patients with *Acinetobacter* spp. infection, with a median LOS of 27 days and 60% of whom were admitted to ITU at some point; furthermore, in those subjects that died there was a prolonged median period after isolation of *Acinetobacter* spp. until death of 16.5 days. Overall, our results provide a clearer clinical picture and better definition of the burden of *Acinetobacter* spp. infections managed in Thai secondary care services.

The sites chosen for our study were secondary care hospitals situated in medium sized towns within largely rural regions of Northern (Lamphun) and Northeastern (Nakhon Phanom) Thailand. Although the lower case numbers at Lamphun did not allow power for formal comparisons between the two sites, in general there were limited observed differences between the two sites. Cases at both sites had similar mean ages, a male predominance, were dominated by respiratory tract infections, and were associated with high rates of ITU admission and mortality. However, there were also some differences between the sites. Reinforcing our previous findings, *A. nosocomialis* infections were associated with high levels of AMR and mortality (23), but interestingly were restricted to the Nakhon Phanom site. In addition, co-infection was commoner at Nakhon Phanom, and the overall rate of weekly infections was twice as high as Lamphun. The reasons behind the differences between the two sites are not clear but presumably relate to the relative exposure to these pathogens at the different geographical sites.

Our data showed that *Acinetobacter* spp. infections were commoner in the rainy season at the Nakhon Phanom site. Seasonal differences in *Acinetobacter* spp. infections is well recognised but previously has largely been described as an increase in infections during warmer seasons (34–36), whereas the Thai rainy season is mainly characterised by increased humidity rather than temperature. An association of *A. baumannii* community-acquired pneumonia (CAP) with the rainy season was suggested for Queensland and Reunion Island (37,38), but not in a previous study from Northeastern Thailand (39). Potential reasons underpinning an increase in *Acinetobacter* spp. infections in the rainy season could be increased exposure to *Acinetobacter* spp. sources of infection, improved transmissibility, or perhaps direct effects on bacterial infectivity. Further investigation into why the rainy season is associated with an increase in *Acinetobacter* spp. infections could also provide important information on what factors underpin the overall high incidence of *Acinetobacter* spp. infections in Thailand.

Another important observation was the high frequency of co-infection with other potential pathogens in respiratory, pus, or blood samples containing *Acinetobacter* spp. Similarly, high rates of co-infection were seen in respiratory isolates in a large Italian study (40). Pus samples had a particularly high rate of infection with multiple pathogens, and this is likely to reflect true mixed infections. The clinical relevance of co-infection in respiratory samples is less clear. Almost all the potential pathogens found in sputum along with *Acinetobacter* spp. cause acute lung infections but are also common commensal species; hence co-infection could reflect dual infection, or active infection with either *Acinetobacter* spp. alone or the co-infecting pathogen alone. The major changes in antibiotic regimens after identification of *Acinetobacter* spp. illustrates the complexities of treating this pathogen caused by its high levels of antibiotic resistance. Co-infection would make identifying an effective antibiotic regimen even more difficult due to the need to target all the infecting pathogens with their potentially different antibiotic resistance patterns.

Despite multiple case series describing *Acinetobacter* spp. CAI (14,15,18–22), these remain an under-appreciated source of infection. At both Lamphun and Nakhon Phanom 25% of infections were CAIs, demonstrating that in Thai secondary care centres CAIs are a major contributor to the high prevalence of *A. baumannii* infections. *A. baumannii* CAIs were frequently severe, with a median LOS of 15 days and a hospital mortality of 18%; strikingly 33% were admitted to ITU on the day of admission. *A. baumannii* CAIs were not restricted to the respiratory tract, with higher incidence of isolation from pus, urine, and blood for CAIs compared to HAIs. This relatively high prevalence and range of *Acinetobacter* spp. CAIs and their lower rates of AMR compared to HAIs indicate that there is a significant source of infection in the community, but this remains uncharacterised. Identifying the source of *Acinetobacter* spp. CAIs would be important, as this could lead to effective preventative measures. Whether *Acinetobacter* spp. CAIs or HAIs represent cross-infection of community and hospital environments or independent sources of infection remains unknown.

This study has several important limitations. Due to the limited research capacity at our research sites, only 20-25% of cases of *Acinetobacter* spp. infection were recruited for the study potentially leading to bias in the data. However, the demographic features and breakdown of sites of infection were largely similar between recruited subjects and all cases of *Acinetobacter* spp. infection, providing reassurance the recruited subjects were not a highly selected population. Some data were incompletely collected, specifically the 90 day mortality data. At both hospital sites patients have a preference to die at home when further treatment was deemed futile and many recruited subjects were lost to follow up after hospital discharge, hence the 90 day mortality should be considered a minimal figure. For patients already on ITU, isolating *Acinetobacte*r spp. from a respiratory sample could reflect colonisation or active infection. In our study 31% of these cases were associated with new consolidation on the chest X-ray suggesting active infection, but whether the remaining subjects had active infection or just colonisation remains unclear. Lastly, the study lacks statistical power to confirm potentially important differences between subgroups; for example, with sample sizes of 96 HAI and 33 CAI cases this study is powered to detect only >50% differences between these groups and then only for parameters that affect at least 60% of the HAI group. Our data support a need for a larger clinical dataset and continued clinical research to fully understand the patient population at risk in this and similar settings, with support to local sites to harmonise and collate data for analysis of clinical characteristics and patient outcomes.

## Conclusions

Our prospective clinical study at two general hospitals in Thailand has confirmed there is a very high burden of *Acinetobacter* spp. infections, the majority of which had high levels of AMR. Although there were a wide range of clinical presentations, *Acinetobacter* spp. were predominately associated with the respiratory tract, causing both CAP and HAIs. A quarter of infections were CAI, emphasising the potential importance of the community as a source of infection. *Acinetobacter* spp. infections led to prolonged hospital stays and a high proportion of ITU admissions and deaths. Overall, these data highlight where to focus data collection required for planning future clinical trials of novel therapeutic approaches that can ameliorate the burden of disease in highly affected countries, and potentially to help identify why there is such a high burden of *Acinetobacter* spp. infection in Thailand.

## Data Availability

All data produced in the present study are available upon reasonable request to the authors.

## Acknowledgments / funding

The project was funded by MRC grants MR/R001871/1 and MR/Y008693/1. The work was undertaken at UCLH/UCL who receive funding from the Department of Health’s NIHR Biomedical Research Centre’s funding scheme. Nakhon Phanom Hospital Team: Neulta Phosawang (Research Nurse), Uraiwan Surin and Chidchanok Promkong (Medical Technologists), and Sirinan Ponhiamhan (Pharmacist). Lamphun Hospital team: Sasiprapa Tansuwat and IC nurse team, Piyapong Pinta (Medical Technologist) and Warangkhana Wanla (Pharmacist).

